# Mindfulness-Based Interventions using Artificial Intelligence: A Systematic Review Protocol

**DOI:** 10.1101/2025.06.20.25329981

**Authors:** Ravi Shankar, Fiona Devi, Xu Qian

**Author notes:** **Corresponding Author:** Dr Ravi Shankar; Research and Innovation, Medical Affairs, Alexandra Hospital, Singapore, Email correspondence.

## Abstract

**Background:** Mindfulness-based interventions (MBIs) have gained significant recognition as effective approaches for promoting mental health and well-being. With rapid advancements in artificial intelligence (AI), there is growing interest in leveraging AI technologies to enhance the delivery, personalization, and effectiveness of MBIs. However, the current state of research in this field remains nascent and fragmented, with limited comprehensive understanding of the types of AI technologies being used, their effectiveness in promoting mental health outcomes, and user experiences with these interventions.

**Objectives:** This systematic review aims to comprehensively synthesize existing literature on AI-based MBIs. The primary objectives are to: (1) identify and characterize the types of AI technologies employed in MBIs, including machine learning, natural language processing, computer vision, and other relevant approaches; (2) evaluate the effectiveness of these interventions in promoting mental health and well-being outcomes, such as reducing symptoms of depression, anxiety, and stress, and enhancing overall quality of life; (3) examine the acceptability, feasibility, and user experience of AI-based MBIs; and (4) identify key trends, research gaps, and challenges within the current literature to provide recommendations for future research and practice.

**Methods and Analysis:** This protocol follows PRISMA-P guidelines and has been registered in PROSPERO (CRD42025641273). Eligibility criteria are based on the PICOS framework, including adults (≥18 years) with or without mental health conditions, mindfulness-based interventions incorporating AI technologies, various comparison groups (including no comparison), mental health and well-being outcomes, and diverse study designs including RCTs, non-randomized studies, and qualitative studies. A comprehensive search strategy will be implemented across multiple databases (PubMed, Web of Science, Embase, CINAHL, MEDLINE, The Cochrane Library, PsycINFO, and Scopus) from inception to January 2025, supplemented by manual searches, gray literature searches, and expert consultation. Two independent reviewers will conduct study selection, data extraction, and risk of bias assessment using appropriate tools (RoB 2 for RCTs, ROBINS-I for non-randomized studies, CASP for qualitative studies). A narrative synthesis will be conducted following Cochrane guidance, with meta-analysis considered if studies are sufficiently homogeneous. The GRADE approach will be used to assess certainty of evidence.

**Discussion:** This systematic review will provide the first comprehensive synthesis of evidence on AI-based MBIs, addressing critical gaps in understanding the integration of AI technologies in mindfulness interventions. The findings will inform the development of best practices and guidelines for designing, evaluating, and implementing AI-enhanced MBIs. By identifying the most promising AI technologies and applications, as well as key research gaps and methodological challenges, this review will guide future research priorities and funding allocation. The results will contribute to ongoing dialogue between researchers, practitioners, and policymakers at the intersection of mindfulness, mental health, and AI, ultimately supporting the broader goal of leveraging AI technologies to enhance the accessibility, effectiveness, and personalization of mindfulness-based interventions for mental health and well-being.

## Introduction

Mindfulness, defined as the practice of purposefully paying attention to the present moment with a non-judgmental attitude, has been increasingly recognized as a valuable approach for promoting mental health and well-being [1]. MBIs, such as mindfulness-based stress reduction (MBSR) and mindfulness-based cognitive therapy (MBCT), have been widely studied and shown to be effective in reducing symptoms of anxiety, depression, and stress, as well as improving overall quality of life [2, 3].

The core components of MBIs typically include formal meditation practices, such as body scan, sitting meditation, and mindful movement, as well as informal practices that involve bringing mindful awareness to daily activities [4]. These practices aim to cultivate a state of present-moment awareness, acceptance, and non-judgmental observation of one’s thoughts, emotions, and bodily sensations. Through regular practice, individuals can develop a greater sense of emotional regulation, cognitive flexibility, and self-compassion [5].

The effectiveness of MBIs has been demonstrated across a wide range of populations and settings, including clinical populations with mental health conditions such as depression, anxiety disorders, and substance use disorders [6, 7], as well as non-clinical populations seeking to reduce stress and improve well-being [8]. MBIs have also been adapted for specific populations, such as pregnant women [9], older adults [10], and healthcare professionals [11].

In recent years, there has been a growing interest in leveraging AI technologies to enhance the delivery and effectiveness of MBIs. AI refers to the development of computer systems that can perform tasks that typically require human intelligence, such as learning, problem-solving, and decision-making [12]. AI encompasses various subfields, including machine learning, natural language processing, and computer vision, which have the potential to be applied to MBIs in innovative ways.

Machine learning, a subset of AI that involves training algorithms to learn patterns from data, can be used to personalize MBIs based on individual characteristics and preferences. For example, machine learning algorithms can analyze user data, such as demographic information, psychometric assessments, and engagement patterns, to recommend specific mindfulness exercises or tailor the content and difficulty level of the intervention [13]. This personalization approach can potentially increase the relevance and effectiveness of MBIs by addressing individual needs and preferences.

Natural language processing (NLP), another subfield of AI, focuses on the interaction between computers and human language. NLP techniques can be applied to analyze user-generated text data, such as journal entries, forum posts, or chat conversations, to identify patterns, sentiments, and topics related to mindfulness practice [14]. This analysis can provide valuable insights into the experiences, challenges, and benefits of mindfulness practice, which can inform the design and improvement of MBIs.

Computer vision, which involves training computers to interpret and understand visual information from the world, can be used to analyze facial expressions, body language, and physiological responses during mindfulness practice [15]. For example, computer vision algorithms can detect facial landmarks and classify emotional states, such as calmness or stress, based on facial expressions. This real-time feedback can be used to provide personalized guidance and support during mindfulness practice, enhancing the user experience and promoting adherence.

Despite the promising potential of AI-based MBIs, the current state of research in this field is still nascent and fragmented. There is a lack of a comprehensive understanding of the types of AI technologies being used, their effectiveness in promoting mental health and well-being outcomes, and the user experience and acceptability of these interventions. Furthermore, there are concerns about the ethical implications of using AI in mental health interventions, such as privacy, data security, and algorithmic bias [16].

To address these gaps and concerns, a systematic review of the current literature on AI-based MBIs is needed. This review will provide a comprehensive synthesis of the existing evidence, identify key trends and gaps, and provide recommendations for future research and practice in this emerging field. The findings of this review will have important implications for researchers, practitioners, and policymakers interested in leveraging AI technologies to enhance the delivery and effectiveness of MBIs.

The primary objective of this systematic review is to comprehensively synthesize the existing literature on artificial intelligence (AI)-based mindfulness-based interventions (MBIs). Specifically, the review aims to identify and characterize the types of AI technologies employed in MBIs, including machine learning, natural language processing, computer vision, and other relevant approaches. It also seeks to evaluate the effectiveness of these interventions in promoting mental health and well-being outcomes, such as reducing symptoms of depression, anxiety, and stress, and enhancing overall quality of life. Furthermore, the review will examine the acceptability, feasibility, and user experience of AI-based MBIs, focusing on factors such as user engagement, satisfaction, and perceived benefits and challenges. In addition, it will identify key trends, research gaps, and challenges within the current body of literature, including methodological limitations, the diversity of populations studied, and ethical considerations. Finally, the review will offer recommendations for future research and practice, highlighting directions for innovation, evaluation, and implementation of AI-based MBIs.

## Methods

### Protocol and Registration

This systematic review protocol follows the Preferred Reporting Items for Systematic Review and Meta-Analysis Protocols (PRISMA-P) guidelines [17]. The protocol has been registered in the International Prospective Register of Systematic Reviews (PROSPERO: CRD42025641273). Any deviations from the protocol will be documented and justified in the final review.

### Eligibility Criteria

The eligibility criteria for this systematic review are based on the PICOS (Population, Intervention, Comparison, Outcome, Study Design) framework [18].

### Population

The population of interest includes adults (aged 18 years and above) with or without mental health conditions. No restrictions will be placed on gender, race, ethnicity, or geographical location. Studies that focus on specific populations, such as healthcare professionals, students, or individuals with chronic illnesses, will be included if they meet the other eligibility criteria.

### Intervention

The intervention of interest is mindfulness-based interventions that incorporate artificial intelligence technologies, such as machine learning, natural language processing, or computer vision. The MBIs can be delivered through various formats, such as mobile apps, web-based platforms, virtual reality, or wearable devices. The duration and frequency of the interventions will not be restricted. Studies that involve AI-based components as an adjunct to traditional MBIs will also be included.

### Comparison

The comparison groups in this systematic review may include a range of control conditions to ensure a comprehensive assessment of AI-based mindfulness-based interventions (MBIs). These include inactive control conditions such as waitlist groups, no treatment, or usual care, as well as active control conditions like traditional MBIs without AI components, other psychological interventions, or educational materials. Additionally, studies comparing different types or dosages of AI-based MBIs will be considered. Importantly, the review will also include studies without a comparison group, such as single-arm pre-post studies, as they can offer valuable insights into the feasibility, acceptability, and preliminary outcomes associated with AI-based MBIs.

### Outcomes

The primary outcomes of interest in this systematic review focus on mental health and well-being indicators. These include symptoms of depression, anxiety, and stress, which may be measured through validated self-report questionnaires such as the Patient Health Questionnaire-9 (PHQ-9), Generalized Anxiety Disorder-7 (GAD-7), and Perceived Stress Scale (PSS), or through clinician-administered instruments such as the Hamilton Depression Rating Scale (HAMD) and the Hamilton Anxiety Rating Scale (HAMA). Other primary outcomes include quality of life and well-being, assessed using generic or condition-specific tools like the SF-36 or WHOQOL-BREF, and mindfulness skills, measured via self-report instruments such as the Five Facet Mindfulness Questionnaire (FFMQ) and the Mindful Attention Awareness Scale (MAAS).

Secondary outcomes include measures of acceptability, such as user satisfaction ratings, adherence rates, and qualitative feedback from participants. Feasibility will be evaluated through metrics such as recruitment and retention rates, as well as app usage data. User experience will be assessed using qualitative methods (e.g., interviews, focus groups) and standardized questionnaires like the User Experience Questionnaire (UEQ) and the System Usability Scale (SUS). Finally, the review will also consider any reported adverse events or unintended consequences, based on participant reports or objective monitoring.

### Study Design

The eligible study designs for inclusion in this systematic review comprise a range of empirical research methodologies. These include randomized controlled trials (RCTs), non-randomized controlled trials (NRCTs), and observational studies such as cohort studies, case-control studies, and cross-sectional studies. In addition, qualitative studies that explore user experiences and perceptions of AI-based mindfulness-based interventions (MBIs) are eligible, as are mixed-methods studies that integrate both quantitative and qualitative approaches. Non-empirical studies, including review articles, commentaries, and theoretical papers, will be excluded from the review. However, conference abstracts and dissertations will be considered for inclusion if they contain sufficient information to assess study eligibility and extract relevant data.

### Information Sources and Search Strategy

A comprehensive search strategy will be developed in consultation with a medical librarian to ensure the identification of all relevant studies on AI-based mindfulness-based interventions (MBIs). The search will cover multiple electronic databases from their inception to January 2025. These databases include PubMed, Web of Science, Embase, CINAHL, MEDLINE, The Cochrane Library, PsycINFO, and Scopus. This broad and systematic approach aims to capture a wide range of peer-reviewed literature across disciplines related to healthcare, psychology, technology, and social sciences.

The search strategy will be adapted for each database and will include a combination of keywords and subject headings related to mindfulness, AI, and mental health. An example search string template is:

((“mindfulness” OR “meditation” OR “mindfulness-based intervention*” OR “MBSR” OR “MBCT”) AND (“artificial intelligence” OR “machine learning” OR “deep learning” OR “natural language processing” OR “computer vision” OR “virtual reality” OR “wearable*”) AND (“mental health” OR “depression” OR “anxiety” OR “stress” OR “well-being” OR “quality of life”))

In addition to the electronic database search, we will manually search the reference lists of included studies and relevant review articles to identify any additional eligible studies. We will also search for ongoing or unpublished studies in clinical trial registries, such as ClinicalTrials.gov and the World Health Organization International Clinical Trials Registry Platform.

Furthermore, we will conduct a gray literature search to identify relevant conference proceedings, dissertations, and reports from organizations involved in mindfulness research or AI development. This search will include databases such as OpenGrey, ProQuest Dissertations & Theses, and IEEE Xplore.

Finally, we will contact experts in the field of mindfulness and AI to inquire about any ongoing or unpublished studies that may be relevant to our review.

### Study Selection

The study selection process will be conducted in two stages using Covidence, a web-based systematic review management platform. First, two independent reviewers will screen the titles and abstracts of the identified studies against the eligibility criteria. Studies that clearly do not meet the criteria will be excluded, while those that appear relevant or uncertain will be retained for full-text review. Second, the full-text articles of the remaining studies will be reviewed independently by the two reviewers to determine their final inclusion. Any discrepancies between the reviewers will be resolved through discussion or by consulting a third reviewer.

The study selection process will be documented using a PRISMA flow diagram [19], which will outline the number of studies identified, screened, eligible, and included in the systematic review. Reasons for exclusion at the full-text stage will be recorded and reported.

### Data Extraction

A standardized data extraction form will be developed using Covidence to systematically collect relevant information from all included studies. To ensure its clarity and comprehensiveness, the form will be piloted on a subset of studies before full implementation. The extracted data will include several key domains: (1) study characteristics, such as first author, year of publication, country, study design, sample size, and funding source; (2) participant characteristics, including age, gender, race/ethnicity, mental health status, and comorbidities; (3) intervention characteristics, detailing the type of mindfulness-based intervention (MBI), AI technologies used, duration, frequency, and delivery format; (4) comparison group characteristics, if applicable; (5) outcome measures and time points of assessment; (6) quantitative results, including means, standard deviations, effect sizes, and p-values; (7) qualitative findings, such as key themes and participant quotes; (8) any reported adverse events or unintended consequences; (9) data on acceptability, feasibility, and user experience; and (10) key conclusions and recommendations provided by the study authors. Data extraction will be conducted independently by two reviewers, with discrepancies resolved through discussion or by involving a third reviewer. If any data are missing or unclear, study authors will be contacted via email to obtain clarification.

### Risk of Bias Assessment

The risk of bias in the included studies will be assessed using appropriate tools based on the study design. For RCTs, the Cochrane Risk of Bias 2 (RoB 2) tool [20] will be used. This tool assesses bias across five domains: randomization process, deviations from intended interventions, missing outcome data, measurement of the outcome, and selection of the reported result.

For non-randomized studies, the Risk Of Bias In Non-randomized Studies of Interventions (ROBINS-I) tool [21] will be used. This tool assesses bias across seven domains: confounding, selection of participants, classification of interventions, deviations from intended interventions, missing data, measurement of outcomes, and selection of the reported result.

For qualitative studies, the Critical Appraisal Skills Programme (CASP) Qualitative Checklist [22] will be used. This checklist assesses the methodological quality of qualitative studies across ten domains, including research design, data collection, and analysis.

Two independent reviewers will assess the risk of bias for each included study, and any discrepancies will be resolved through discussion or by consulting a third reviewer. The results of the risk of bias assessment will be presented in a tabular format and will be considered in the interpretation of the review findings.

### Data Synthesis

A narrative synthesis of the included studies will be conducted, following the guidance provided by the Cochrane Handbook for Systematic Reviews of Interventions [23]. The synthesis will be structured around the review objectives and will consider the types of AI technologies used, the effectiveness of AI-based MBIs, the acceptability and feasibility of these interventions, and the user experience.

The narrative synthesis will begin by summarizing the characteristics of the included studies, such as the study designs, populations, interventions, and outcomes. This summary will provide an overview of the evidence base and highlight any notable differences between studies.

Next, we will synthesize the quantitative findings on the effectiveness of AI-based MBIs. For each outcome, we will report the direction and magnitude of the effects, along with the precision and consistency of the findings. We will also consider the clinical significance of the effects and any potential moderating factors, such as participant characteristics or intervention features.

If sufficient data are available and the included studies are homogeneous in terms of interventions and outcomes, a meta-analysis will be considered to quantitatively synthesize the effectiveness of AI-based MBIs. The meta-analysis will be conducted using a random-effects model, as it allows for heterogeneity between studies [24]. Heterogeneity will be assessed using the I^2^ statistic, which describes the percentage of variation across studies that is due to heterogeneity rather than chance [25]. If substantial heterogeneity is detected (I^2^ > 50%), we will explore potential sources of heterogeneity through subgroup analyses or meta-regression, if feasible.

Subgroup analyses may be performed based on participant characteristics (e.g., age, gender, mental health status), intervention types (e.g., machine learning, NLP, computer vision), or study designs (e.g., RCTs vs. NRCTs). These analyses will help to identify any differential effects of AI-based MBIs across different subgroups and inform the development of tailored interventions.

For qualitative findings, we will conduct a thematic synthesis [26] to identify common themes and patterns across studies related to the acceptability, feasibility, and user experience of AI-based MBIs. The synthesis will involve coding the qualitative data, generating descriptive themes, and developing analytical themes that go beyond the primary studies. The qualitative findings will be integrated with the quantitative results to provide a more comprehensive understanding of AI-based MBIs.

If a meta-analysis is not feasible due to heterogeneity or insufficient data, a narrative synthesis will be conducted, focusing on the patterns, similarities, and differences across studies. The synthesis will consider the methodological quality of the studies and the potential impact of bias on the findings.

Throughout the synthesis process, we will assess the certainty of the evidence using the Grading of Recommendations Assessment, Development, and Evaluation (GRADE) approach [27]. The GRADE approach considers factors such as study design, risk of bias, inconsistency, indirectness, imprecision, and publication bias to provide an overall assessment of the certainty of the evidence for each outcome.

### Reporting Bias Assessment

To assess the potential impact of reporting biases, such as publication bias or selective outcome reporting, we will use appropriate methods based on the type of data available.

For quantitative outcomes, if there are at least ten studies included in a meta-analysis, we will create funnel plots to visually assess the presence of asymmetry, which may indicate publication bias [28]. We will also perform statistical tests, such as Egger’s test [29] or Begg’s test [30], to quantitatively assess funnel plot asymmetry. If publication bias is suspected, we will conduct a sensitivity analysis using the trim-and-fill method [31] to estimate the impact of missing studies on the pooled effect size.

For qualitative outcomes, we will consider the potential for selective reporting of themes or participant quotes that support the authors’ conclusions. We will compare the reported themes with the raw data (if available) to assess the completeness and transparency of the qualitative synthesis.

If reporting biases are identified, we will discuss their potential impact on the review findings and consider the implications for the interpretation and generalizability of the results.

## Discussion

This systematic review protocol presents a comprehensive and methodologically rigorous approach to synthesizing the current evidence on artificial intelligence (AI)-based mindfulness-based interventions (MBIs). It aims to make a timely and meaningful contribution to the field by addressing several critical gaps. First, it responds to the rapid growth of AI technologies and their transformative potential in enhancing the delivery, personalization, and effectiveness of MBIs. Second, it highlights the current lack of a thorough understanding of the specific types of AI technologies being integrated into these interventions, their mental health outcomes, and users’ experiences with them. Finally, the review underscores the need for a systematic and critical evaluation of the existing literature to guide future research, inform best practices, and support policy development in the implementation of AI-enhanced mental health interventions.

The strengths of this protocol include its adherence to the PRISMA-P guidelines, the use of a comprehensive search strategy across multiple databases, the inclusion of a wide range of study designs and outcomes, and the planned assessment of risk of bias and certainty of evidence using established tools and frameworks.

However, there are also potential limitations and challenges to consider. The heterogeneity of AI technologies and their applications in MBIs may make it difficult to synthesize the evidence and draw firm conclusions. The rapidly evolving nature of the field may result in the identification of studies using novel or emerging AI approaches that may not be directly comparable to earlier studies.

Furthermore, the inclusion of studies with diverse designs, populations, and outcome measures may introduce additional sources of heterogeneity and complicate the interpretation of the findings. The potential for reporting biases, particularly in the context of a nascent and highly interdisciplinary field, may also impact the validity and reliability of the evidence.

Despite these challenges, this systematic review has the potential to make important contributions to the field of AI-based MBIs. By providing a comprehensive and critical synthesis of the current evidence, the review can help to identify the most promising AI technologies and applications, as well as the key research gaps and methodological issues that need to be addressed in future studies.

The findings of this review can inform the development of best practices and guidelines for the design, evaluation, and implementation of AI-based MBIs. They can also guide the prioritization of research efforts and funding allocation to areas with the greatest potential for impact and innovation.

Furthermore, the review can contribute to the ongoing dialogue and collaboration between researchers, practitioners, and policymakers working at the intersection of mindfulness, mental health, and AI. By bringing together evidence from diverse disciplines and perspectives, the review can foster a more integrated and holistic understanding of the opportunities and challenges of AI-based MBIs.

In conclusion, this systematic review protocol provides a roadmap for a timely and important synthesis of the evidence on AI-based MBIs. The review has the potential to advance the field by providing a comprehensive and critical evaluation of the current state of the evidence, identifying key research gaps and priorities, and informing the development of best practices and guidelines for the design and implementation of AI-based MBIs. The findings of this review can ultimately contribute to the broader goal of leveraging AI technologies to enhance the accessibility, effectiveness, and personalization of mindfulness-based interventions for mental health and well-being.

## Data Availability

This is a systematic review protocol. No primary data were collected for this study. The protocol has been registered in PROSPERO (CRD42025641273). Upon completion of the systematic review, all extracted data will be made available upon reasonable request to the authors.

https://www.crd.york.ac.uk/PROSPERO/view/CRD42025641273

